# Signed Distance Correlation (SiDCo): A network analysis application of distance correlation for identifying metabolic networks disrupted in Dementia with Lewy Bodies

**DOI:** 10.1101/2021.10.16.21265003

**Authors:** Miroslava Čuperlović-Culf, Ali Yilmaz, David Stewart, Anuradha Surendra, Sumeyya Akyol, Sangeetha Vishweswaraiah, Xiaojian Shao, Irina Alecu, Thao Nguyen-Tran, Bernadette McGuinness, Peter Passmore, Patrick G. Kehoe, Michael E. Maddens, Brian D. Green, Stewart F. Graham, Steffany A.L. Bennett

## Abstract

**Motivation:** Identifying pathological metabolic changes in complex disease such as Dementia with Lewy Bodies (DLB) requires a deep understanding of functional modifications in the context of metabolic networks. Network determination and analysis from metabolomics and lipidomics data remains a major challenge due to sparse experimental coverage, a variety of different functional relationships between metabolites and lipids, and only sporadically described reaction networks.

**Results:** Distance correlation, measuring linear and non-linear dependences between variables as well as correlation between vectors of different lengths, e.g. different sample sizes, is presented as an approach for data-driven metabolic network development. Additionally, novel approaches for the analysis of changes in pair-wise correlation as well as overall correlations for metabolites in different conditions are introduced and demonstrated on DLB data. Distance correlation and signed distance correlation was utilized to determine metabolic network in brain in DLB patients and matching controls and results for the two groups are compared in order to identify metabolites with the largest functional change in their network in the disease state. Novel correlation network analysis showed alterations in the metabolic network in DLB brains relative to the controls, with the largest differences observed in *O*-phosphocholine, fructose, propylene-glycol, pantothenate, thereby providing novel insights into DLB pathology only made apparent through network investigation with presented methods.

## 1 Introduction

Elucidating metabolic changes that associate with risk and resiliency in neurodegenerative disease represents a new direction in dementia research, one aimed at targeting the metabolic processes required for preclinical patients to exhibit cognitive symptoms (Alecu and Bennett, 2019; Foolad, et al., 2019; Hallett, et al., 2019). This approach requires analyses of biological networks as a parallel investigation to individual feature characteristics and establishing the interconnection of these features within a biological system while comparing different disease conditions (Ma’ayan, 2011). Methods for such biological network development are broadly divided into knowledge-driven and data-driven approaches where knowledge-based networks primarily aim to contribute to data interpretation while data-based networks derive novel interactions from the data (Amara, et al., 2022). Combining these two approaches can help identify biologically active processes that associate with disease progression. Network analysis is particularly important in metabolomics where extensive maps of all possible metabolic connections are available; however, the active components of the network are specific to the state of the biological system under investigation and have to be obtained from the system data.

Several methods for data-driven network determination have been used in metabolomics. Pearson or Spearman correlation-based methods are arguably the most prevalent (Amara, et al., 2022). While providing critical information about the direction of dependencies, both methods measure linear or monotonic correlations and cannot detect non-linear metabolite interactions (Rosato, et al., 2018). Other correlation-based methods, such as weighted gene correlation network analysis (WCGNA) are widely used to interrogate transcriptomic data (Langfelder and Horvath, 2008). WCGNA has only begun to be applied to metabolomics (Grapov, et al., 2015; Pei, et al., 2017). The main advantage of WCGNA is the capacity to determine the threshold value automatically by assuming a scalefree network topology (Langfelder and Horvath, 2007). It is important to note that the assumption of a scale-free network structure does not necessarily apply to metabolic networks resulting in information loss (Lee, et al., 2008). Distance correlation, a novel non-parametric approach for correlation analysis, has been proposed as a measure of all types of data relationships (linear and non-linear) as well as correlations between vectors of different lengths (Edelmann, et al., 2021; Gábor and Maria, 2009; Székely and Rizzo, 2013; Székely and Rizzo, 2013). Distance correlation can take into consideration the sparse coverage of metabolomic and lipidomic data, the potential for non-linear relationships, the non-normal distribution as well as the possibly random network topology associated with metabolism; however, it cannot describe the direction of the dependencies. While under investigation for general characteristics (Edelmann, et al., 2021; Shen and Zhang, 2021) and with several publications showing its use in transcriptomic assessments (Hou, et al., 2022; Pardo-Diaz, et al., 2021), only a handful of applications have applied this method to analysis of metabolomic datasets (Cuperlovic-Culf, et al., 2021; Oliveira, et al., 2015; Tang, et al., 2019) and none, to our knowledge, have utilized this approach to provide insight into the metabolic network disruptions that define neurodegenerative diseases such as dementias.

Taken together, issues of missing metabolic reactions, sparse analytical coverage, different numbers of measurements for different molecules, and the potential for non-linear or indirect metabolite relationships emphasizes the need for further development and application of novel methodologies applicable to metabolomics. To address this need, we compare here the metabolic networks elucidated by Pearson, distance, and a novel hybrid signed distance correlation approach in two metabolomic datasets of post-mortem human brain of cognitively normal controls and individuals with Dementia with Lewy Bodies (Akyol, et al., 2020). We show here that Pearson correlation fails to observe number of significant relationships that are readily obtained by distance correlation calculations. Additionally we provide approaches for selection of major differences between correlation levels both for individual, pair-wise correlations and pan-metabolite wide changes. Using metabolomics and lipidomics data for DLB we show that this novel approach for correlation analysis combined with novel methods for comparison between networks provides highly relevant information about the metabolic changes in the disease state. SidCo (Signed distance correlation) method is provided as a Python package and Web-based application (https://complimet.ca/sidco).

## 2 Methods

### 2.1 Sample collection and datasets

Sample collection and patient characteristics are described in detail in (Akyol, et al., 2020). Briefly, the neocortex (Brodmann area 7) was collected from patients with histopathologically confirmed DLB (n=15) or from ageand sexmatched controls with no known neurological disease (n=30). Tissues were obtained from the Brains for Dementia Research Group, Institute of Clinical Neurosciences, School of Clinical Sciences, University of Bristol, Bristol, UK as part of the study approved by the Beaumont Health System’s Human Investigation Committee (HIC No.: 2018-387), following all approved guidelines. The acquisition, normalization, and quantification of metabolite concentrations by ^1^H NMR and high performance liquid chromatography electrospray ionization tandem mass spectrometry are described in detail in (Akyol, et al., 2020). Complete dataset comprising 215 analytes (log-normalized concentrations) is provided as Supplementary Table 1. Patient demographics information is outlined in Box 1.

#### Box 1.

Patient demographics information

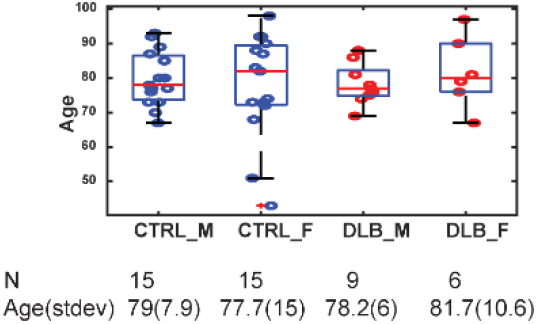

### 2.2 Overview of SidCo data analysis

The SiDCo workflow is presented in Figure 1. Prior to analysis, the user defines the array to be analyzed, the distance correlation, and the p-value thresholds. Data are automatically z-score normalized across all samples. Distance correlations and p-values are calculated as described below and a directionality sign is assigned using Pearson correlation as an indication of the overall trend in the data. It is important to note that the strength of the correlation coefficient does not indicate the strength of the linear correlation, but rather a strength of distance correlation, with sign considering overall linear trend. The output file contains both the correlation coefficients and the p values for each calculation. Distance correlation calculations in SiDCo are provided in two forms “one-to-one”, calculating correlations between each pair of features and “one-to-all”, providing correlations for each feature with all the other features combined. The signed distance correlation calculation is provided as a Web application at http://complimet.ca/SiDCo with a RShiny front-end GUI interface for a Python implementation of the method which includes automatic z-score normalization and signed distance correlation calculation.

**Figure 1.**
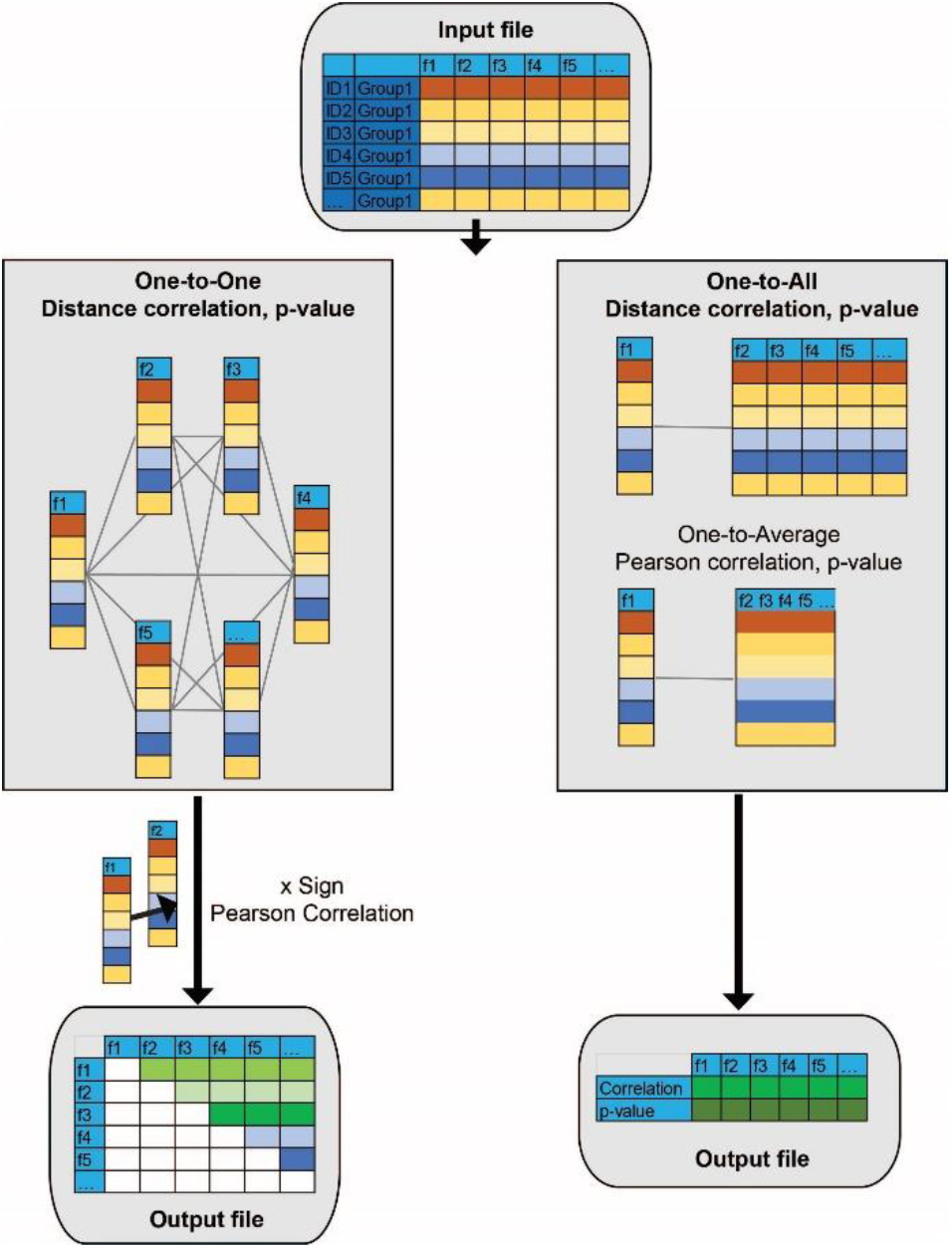
Outline of the SIDCO software for signed distance correlation analysis in one-to-one and one-to-all mode.

In both one-to-one and one-to-all calculations distance correlation is obtained following formalism presented by Szekely and Rizzo (Edelmann, et al., 2021) as defined in Eq. 1 using in-house developed Matlab functions and Python implementation. Distance correlation, *dCor(X, Y*) between features *X* and *Y* is calculated as:

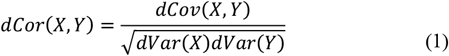

In contrast to Pearson correlation which uses covariance between values obtained as: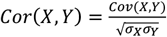, with covariance calculated as: 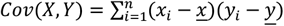, Distance correlation depends on the distance covariances that are determined as:

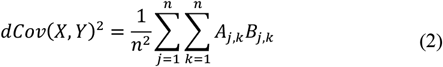

with *A* and *B* calculated as simple linear functions of the pairwise distances between elements in samples *X* and *Y. A* and *B* are doubly centered distance matrices for variables *X* and *Y*, respectively, calculated from the pairwise distance between elements in each sample set. Slight differences in calculating *A* and *B* in one-to-one and one-to-all applications are outlined.

### Distance correlation one-to-one calculation

In one-to-one calculation array of values for each feature is compared with array of values for all other feature one at a time. In this comparison two feature arrays have the same length and are one dimensional arrays.

In this case doubly centered distance matrix is calculated using:

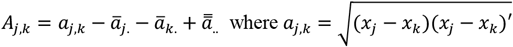

and ā_*i*._ and ā_*k*._ are respectively the *j*-row and *k*-column mean values and 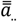. is the overall mean of matrix *A*. Distance between *x*_*j*_ and *x*_*k*_ is calculated using Euclidian distance. Matrix *B* is populated using equivalent measures for variable *Y*. The distance correlation calculation was written in-house under Matlab using *pdist2* to calculate distances between features with Euclidean metric and implemented in Python in SiDCo (https://complimet.ca/sidco). For distance correlation *p*-value is calculated using Student’s *t* cumulative distribution function (*tcdf* function in Matlab and *t*.*cdf* in Python). The sign of the distance correlation was equated to the sign of the Pearson correlation calculation as shown by (Pardo-Diaz, et al., 2021) and implemented in SiDCo as:

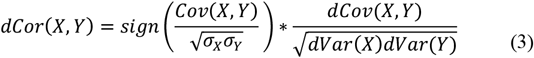

### Distance correlation one-to-all calculation

Additionally, SiDCo offers distance correlation between each feature and all the other features (denoted as one-to-all or the correlation of a given feature with the entire network). Here, the distance covariance for each feature out of *M* features in *N* dimensional sample space is compared to that of the other features in *N* x (*M*-1) dimensional space. Distance correlation and distance covariance are calculated using Equation (1) and (2) where *X* is in this case array of values for one feature across all samples (*N* x1 array) and *Y* is matrix of all other features across all samples (*N* x (*M*-1) matrix). Doubly centered distance matrix for variable *Y* is calculated as:

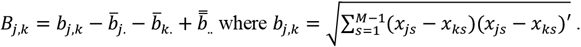

Calculation is in this case performed for each feature correlated to all the other features. SiDCo provides distance correlation value as well as *p*value. In Matlab library SiDCo *p*-value is once again calculated using student cumulative distribution function (function *tcdf*). In Python one-to-all distance correlation is performed using pingouin.distance_corr function (running under pingouin statistical package: https://pingouin-stats.org/index.html) with *p*-value evaluated using permutation test as described in the original application documentation.

Detailed instructions and examples of use for SiDCo are provided on the website.

### Correlation coefficient comparison methods

Two different approaches for comparison of correlations between different sample groups are presented in this work.

#### Box 2.

Example of linear regression analysis approach for selection of features showing major difference between correlation measures for two groups. Negative slope would indicate major change in the overall network for a feature.

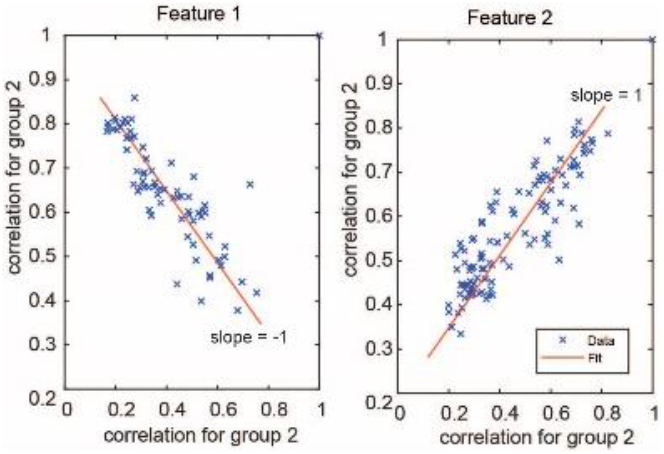

Comparison between individual correlation coefficients obtained between pairs of features using either different methodologies or different sample groups is performed using Fisher z-transformation. Fisher z-transform changes sampling distribution of correlation coefficients into normal distribution for statistical analysis. Fisher z-transformed values are calculated as: 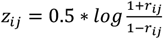 where *r*_*ij*_ is the correlation coefficient between features *i* and *j*. Difference between correlation values is obtained as:

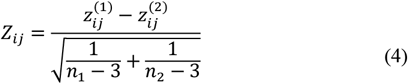

where, respectively, 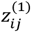 and 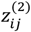are Fisher z-transformed correlation values for group 1 with *n*_*1*_samples and group 2 with *n*_*2*_ samples used for correlation calculations and 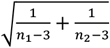 is standard deviation for two sample sets. Statistical significance of the obtained difference between correlation values is calculated using normal cumulative distribution function calculation following Z matrix normalization and analysis of normal distribution with mean 0 and standard deviation of 1. Significant difference between correlation values is associated with p<0.05.

Overall changes in the correlation network for each feature between different methods or sample groups is performed through linear regression comparison of two correlation groups. By optimizing linear function between correlation calculations for a feature obtained for one group using data from the second group we determine overall changes in correlations. Demonstration of this approach is shown in schematic box 1.

Slope values that deviates significantly from 1 indicates major change in overall correlation network between compared conditions. Linear regression obtains parameters for prediction of one variable using the other variable with possible differences in result depending on the dependent and independent group assignment.

To compare approaches, Pearson, distance, and signed distance correlation values with *p*>0.05 were set to zero in subsequent analyses, keeping only the most statistically significant correlations. Correlation results are presented using circular plots created with circularGraph [https://github.com/paul-kassebaum-mathworks/circularGraph] with some in-house modifications.

Matlab functions for distance correlation, Fisher z-transformation and significance analysis as well as correlation linear regression analysis are provided on GitHub.

## 3 Results

### General sample characteristics

Both ^1^H NMR and LC-ESI-MS/MS metabolomics were used to collect information about the concentration of 215 metabolites in brain samples from DLB patients and age and sex-matched healthy controls. Concentrations are reported in Supplementary Table 1. Sample information is summarized in Figure 2A. Principal Component Analysis (PCA) of log-corrected and z-score normalized brain metabolomics data showed some limited grouping of samples by disease diagnosis in PC1 but no clear unsupervised separation by sample type (Figure 2B). This is further corroborated with Wilcox Rank Sum test showing that PC1 provides separation by diagnosis and both PC1 and PC2 are statistically significantly related to subjects’ age (Table in Figure 2B). Similarly, hierarchical clustering performed on z-score normalized data and using Ward linkage indicated clustering by molecular groups but, similarly to PCA, not explicit grouping by sample type (Figure 2C). Relieff (Lionelle, 2005) selected top four features including: O-Phosphocholine, sn-glycerol-3-phosphate, PC(O34:0) and Putrescine provide an improvement in the separation of DLB and CTRL subjects although with several DLB patients still clustering within the control group (Figure 2B).

**Figure 2.**
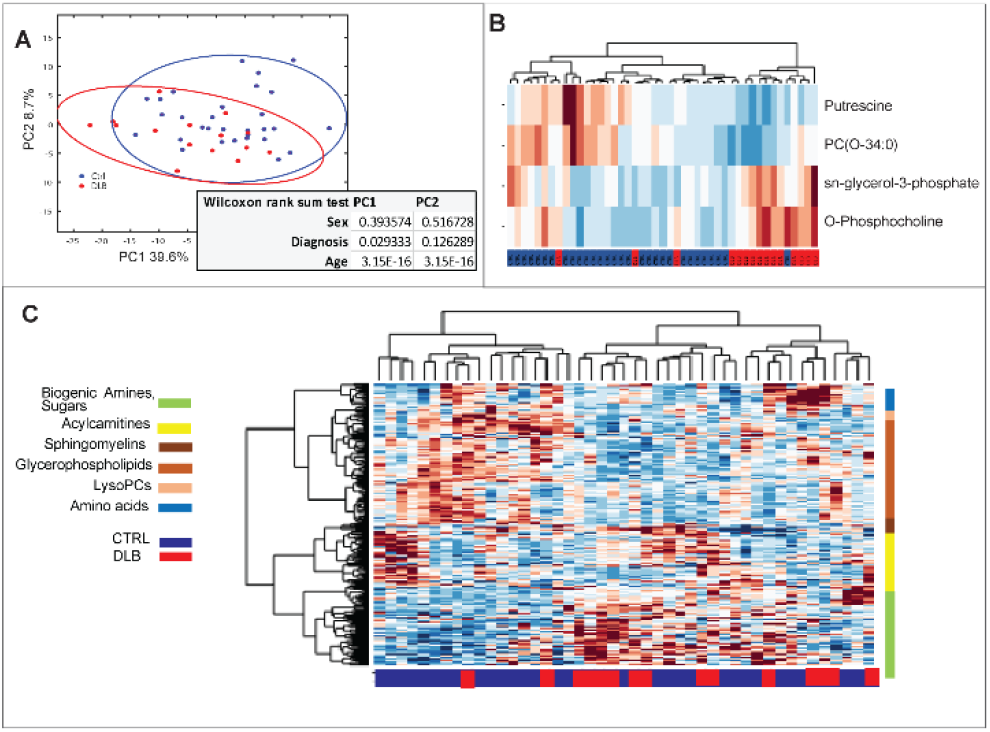
(A) Principal Component Analysis (PCA) of log transformed and z-score normalized features (metabolite levels). Ellipses indicate 95% confidence regions. (B) The most significantly different metabolites between control and DLB selected by Relieff (C) Hierarchical clustering (HCL) of samples and features following z-score normalization of features, with colours on the right indicating groups of metabolites and colours along the bottom indicating control and DLB samples.

### Comparison of metabolic networks derived from pair-wise Pearson correlation and distance correlation

A data-driven network of features in the two groups was determined using Pearson, distance correlation, and signed distance correlation calculation, building on (Székely and Rizzo, 2013). In latter assessments, both proximal as well as distant interactions, including both non-linear and linear correlations, were established. Combining distance correlation with Pearson’s correlation (see Materials and Methods) provided signed distance correlation values indicating both strength and overall directionality of correlations.

Fisher z-transformation of correlation data was used to normalize the distribution of correlation values in order to allow determination of the significance and the difference in correlation values obtained using Pearson and distance methods in control and DLB groups (Fig. 3). Generally, and not surprisingly, for all feature pairs distance correlation leads to higher values with subset of correlation pairs showing statistically significant difference between the two methods. Supplementary Figure 1A. provides the percentage of statistically different correlations for each feature between the two methods in each sample groups. The largest number of statistically significant differences are observed for symmetric dimethyl arginine (SDMA) and spermine in CTRL and PC(26:0) in DLB and scatter plot of results from two correlation methods for these three metabolites are shown (Supplementary Figure 1B). It is important to observe in these examples several metabolite pairs that have significant distance correlation and zero Pearson correlation including mutual correlations between SDMA, urea, fumarate and spermine all metabolites that are part of urea cycle and thus can be expected to show some level of correlation that is obtained by distance correlation but omitted in linear analysis with Pearson correlation.

**Figure 3.**
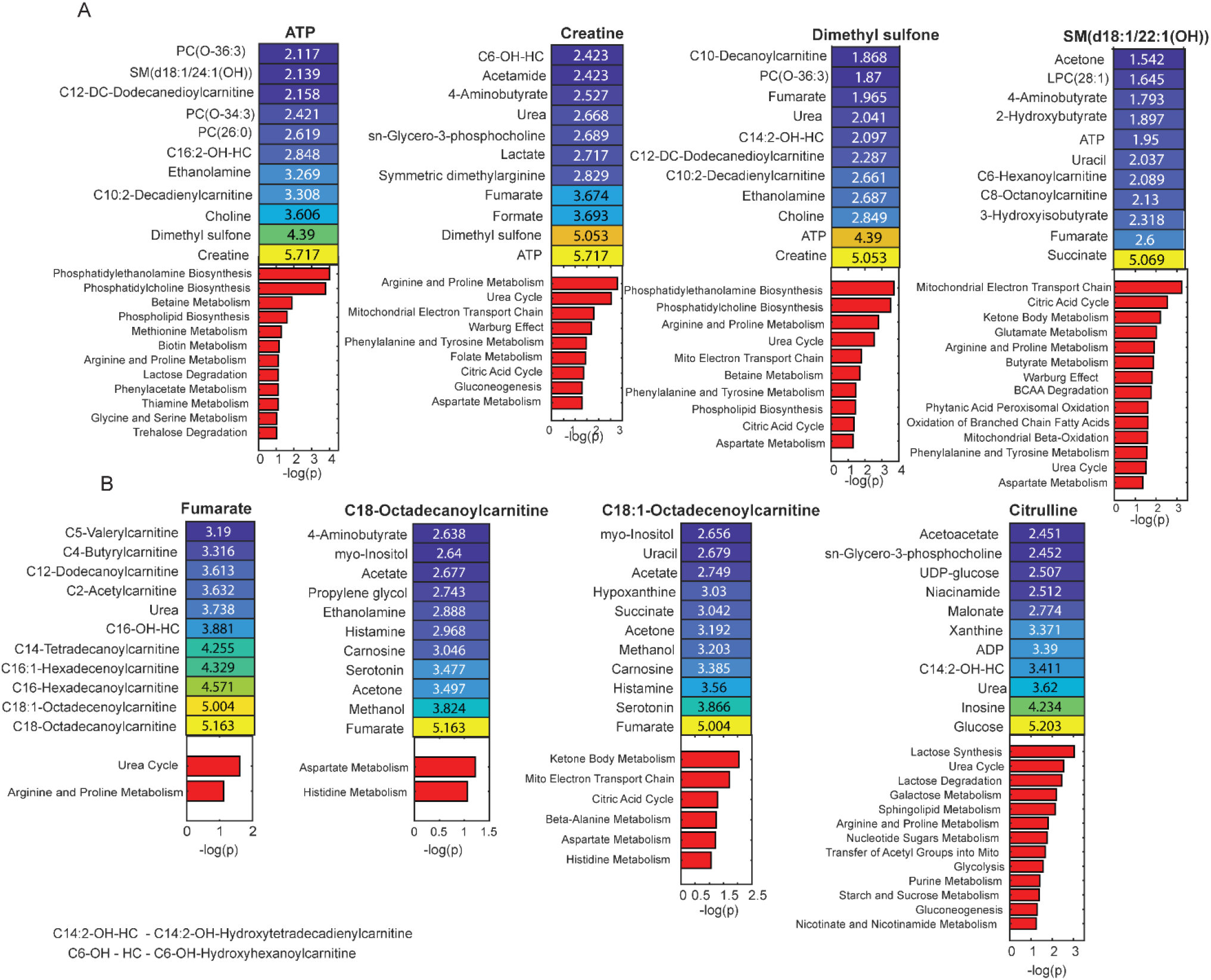
Metabolite pairs with the highest differences between Fisher ztransformed Pearson and distance correlations for A. control and B. DLB groups. Shown are metabolite pairs and correlation difference (heatmap) and enrichment analysis result for these metabolites obtained using Metaboanalyst (red bar plot with –log(p) values for enrichment analysis are indicated).

Further investigation focused on the largest differences between correlation values of Fisher z-transformed Pearson and distance correlation in control (Figure 4A) and DLB groups (Figure 4B). In this case it is observed that number of metabolite pairs have zero or extremely low Pearson correlations and non-zero distance correlations. Shown are differences for the correlation levels for metabolites that have the largest pair-wise distance in standard error units of over 5. For these metabolites and their largest pair-wise correlation distance metabolic partners we have performed pathway enrichment assessment. Enrichment analysis of these metabolite groups shows significant representation of processes known to be of significance in the neurodegeneration and dementia indicating that linear correlation observed with Pearson method is insufficient to provide information about many relevant aspects of the correlation network. Presented examples of correlations that were not observed in Pearson’s analysis show advantage of distance correlation approach for possibly complex, non-linear and indirect relationships crucially important for metabolomics data analysis including processes of relevance in neurodegeneration.

**Figure 4.**
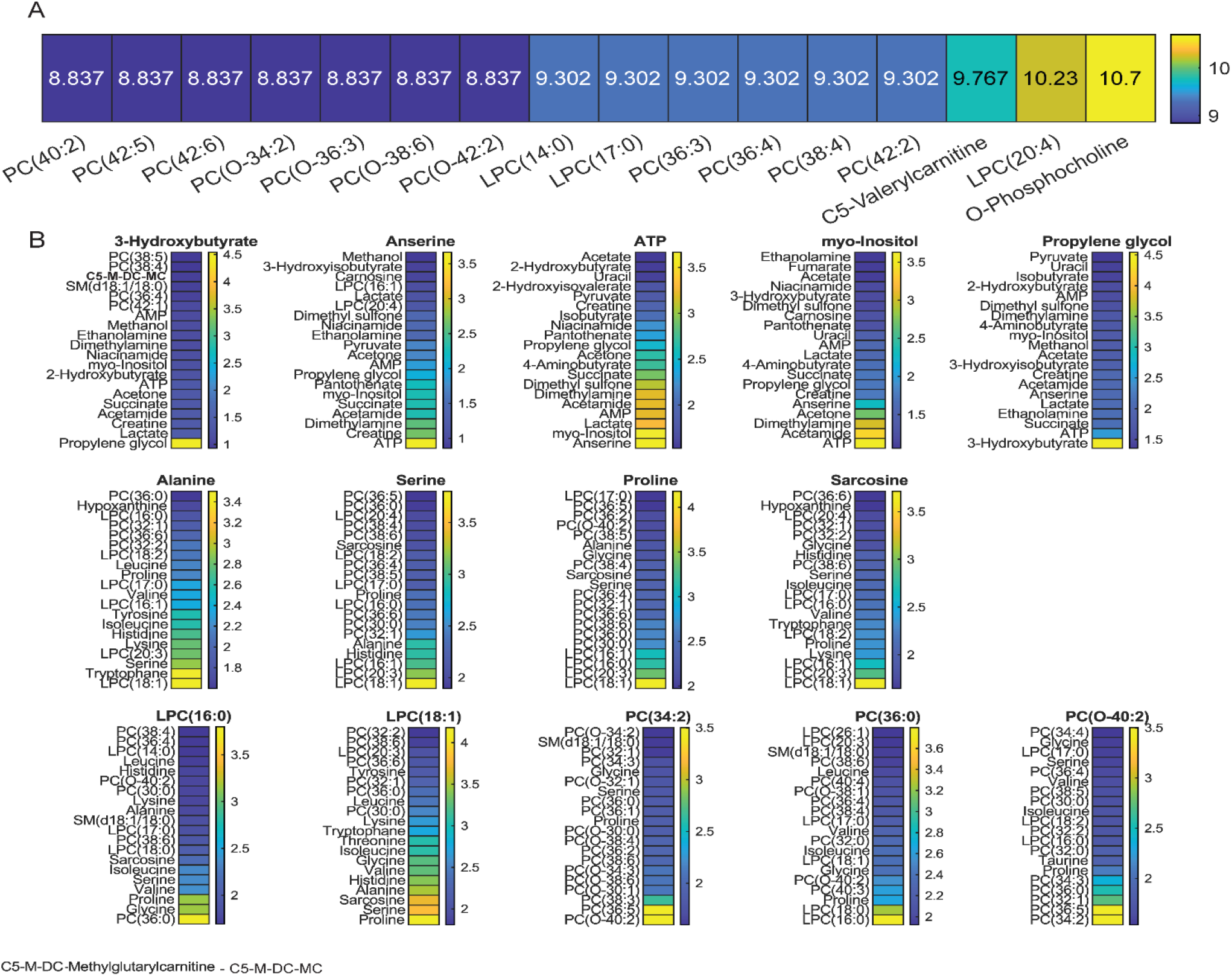
Statistical comparison of the Fisher z-transformed distance correlation values between control and DLB. A. Percentage of pairs with statistically significant change in distance correlation value in two sample groups. Shown are metabolites with the largest number of changed partners. B. Metabolites with the largest difference in pair-wise correlation and their other 15 metabolic partners with the largest change in correlation. Differences are calculated using equation 4.

### Distance correlation analysis as a preferred method for determination of active metabolic interactions

Fisher z-transformation is used for comparing distance correlation values between patient groups. Assessment of normality of the distance correlation values is done using Lilliefors test running under in-house routine for distance correlation calculation. For all correlations in both control and DLB groups null hypothesis that the transformed correlation for each feature to all the other features comes from a normal distribution was shown as significant with p-value approaching zero in all cases. Following this confirmation we have performed analysis of both statistical significance of distance correlation change as well as analysis of the level of change in Fisher z-transformed data and major results are shown in Figure 4. Analysis of significantly changed correlations between two groups shows that *O*-phosphocholine, that was also selected as the species with the largest concentration difference between groups (Figure 2D) is a feature with the largest number of significant changes between control and DLB groups (Figure 4A). *O*-phosphocholine is in this analysis followed by a number of phospholipids all showing statistically significant change in their correlation levels in control and DLB groups. Overall, out of 22898 possible correlation pairs, 1642 show highly statistically significant difference between control and DLB group (p<0.01)s. Further major changes in the level of pair-wise correlation in control and DLB groups are explored with Figure 4B showing species with the largest maximal pair-wise correlation change between two sample groups and lists their other most changed correlation partnerships. Once again, many of the species showing the single largest pair-wise correlation changes are phospholipids with particularly significant change observed in correlations between number of amino acids and phospholipids. This is in agreement with previously determined role of amino acids and particularly serine in phospholipid synthesis (Hirabayashi and Furuya, 2008). Correlation networks in the two sample groups for *O*-phosphocholine and serine, as metabolites with major changes presented below are shown in Figure 7.

An alternative approach to selection of functionally altering major feature compares overall correlation network for each metabolite between two sample groups. In this case comparison is on the original statistically significant distance correlation levels in the two groups using linear regression comparison as described in Materials and Methods. Metabolites showing the most significant change based on this comparison are outlined in Figure 5 with Figure 5B showing scatter plot of the correlation values in the two sample groups for selected metabolites as well as linear regression plot. Figure 5A displays the major network differences obtained from the linear regression analysis of 1-to-1 metabolite correlations in DLB vs. control. Figure 5B shows the value of the slope of the linear regression plot showing control values as a function of distance correlation values in DLB (see Materials and Methods). In this analysis, fructose shows the largest change in its pairwise metabolic correlation network between DLB and control followed by prolylene glycol, SM(d18:1/20:2) and panthoneate. Correlation network nearest partners for these four metabolites in control and DLB groups are shown in Figure 7A and B respectively.

**Figure 5.**
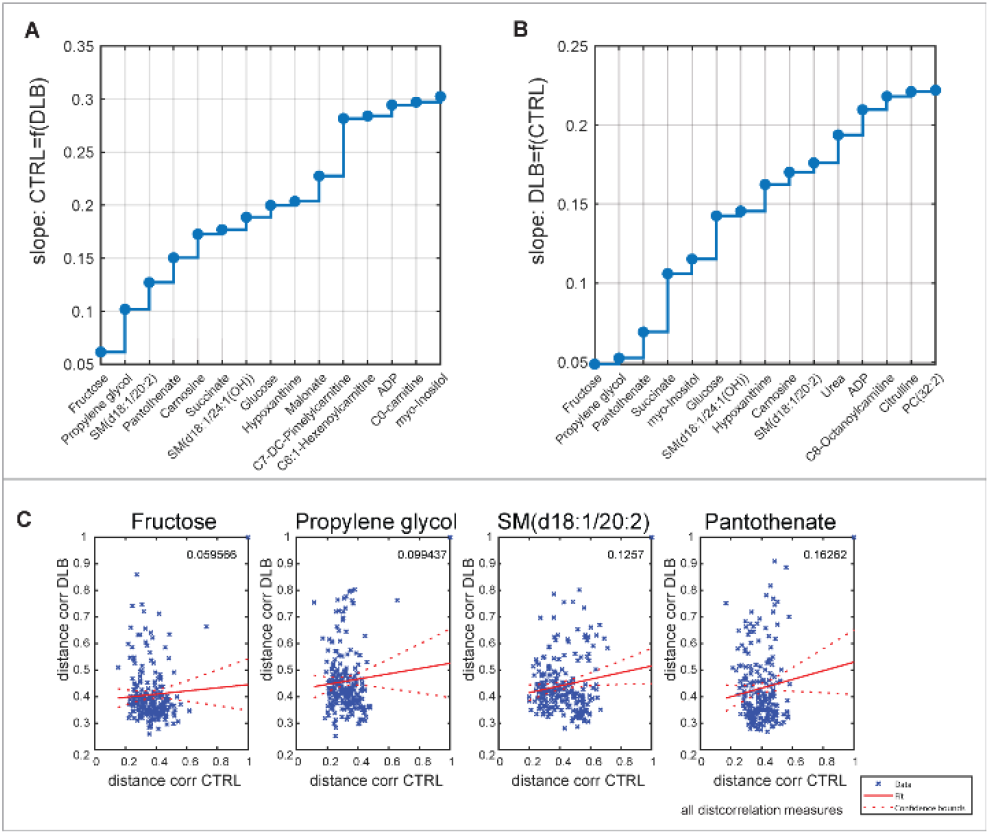
Comparison of Distance correlation network between control and DLB using linear regression comparison (A) metabolites with the largest deviation from slope of 1 in linear regression analysis of overall correlation differences for each metabolite (B) Examples of linear regression result for metabolites with the largest difference from the slope 1.

It is important to stress out that apart from *O*-phosphocholine none of the metabolites selected through distance correlation analysis show statistically significant differences in concentrations between the control and DLB groups (Figure 6); however, their correlation values and main partners change, suggesting the possibility that different pathways are activated for these metabolites in the DLB group.

**Figure 6.**
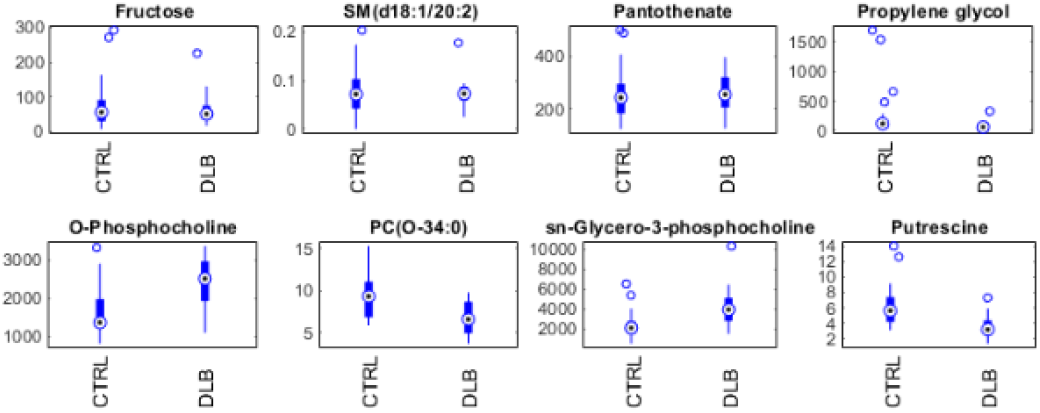
Concentrations of metabolites selected by network analysis and Relieff as significant in control and DLB.

Based on the distance correlation network analysis (Figure 7), the fructose network changes from only a minimal number of significant correlations in the control group to a much larger number of significant edges in the DLB group, leading to a hypothesize that fructose plays a more significant role in the brains of DLB patients. Similarly, the second and third most significantly altered metabolic networks (propylene-glycol and pantothenate metabolism) show more correlations with other metabolites in the DLB cohort. To the best of our knowledge, endogenous propyleneglycol has not been linked to dementia previously, possibly due to its limited concentration change. However, several pathways related to propylene-glycol have been reported as associated with dementia (Killingsworth, et al., 2021). On the other hand, pantothenate, i.e. vitamin B5, has been previously observed to be significantly changed in different types of dementias (Andres-Hernando, et al., 2019; Xu, et al., 2016). Similarly to fructose, based on distance correlation network analysis, pantothenate goes from having no significant correlations in the control group to a number of significant correlations in the DLB group. Strong correlation partners of pantothenate include a metabolites that show significant concentration changes (Figure 2) as well changes in correlation network (Figure 4).

**Figure 7.**
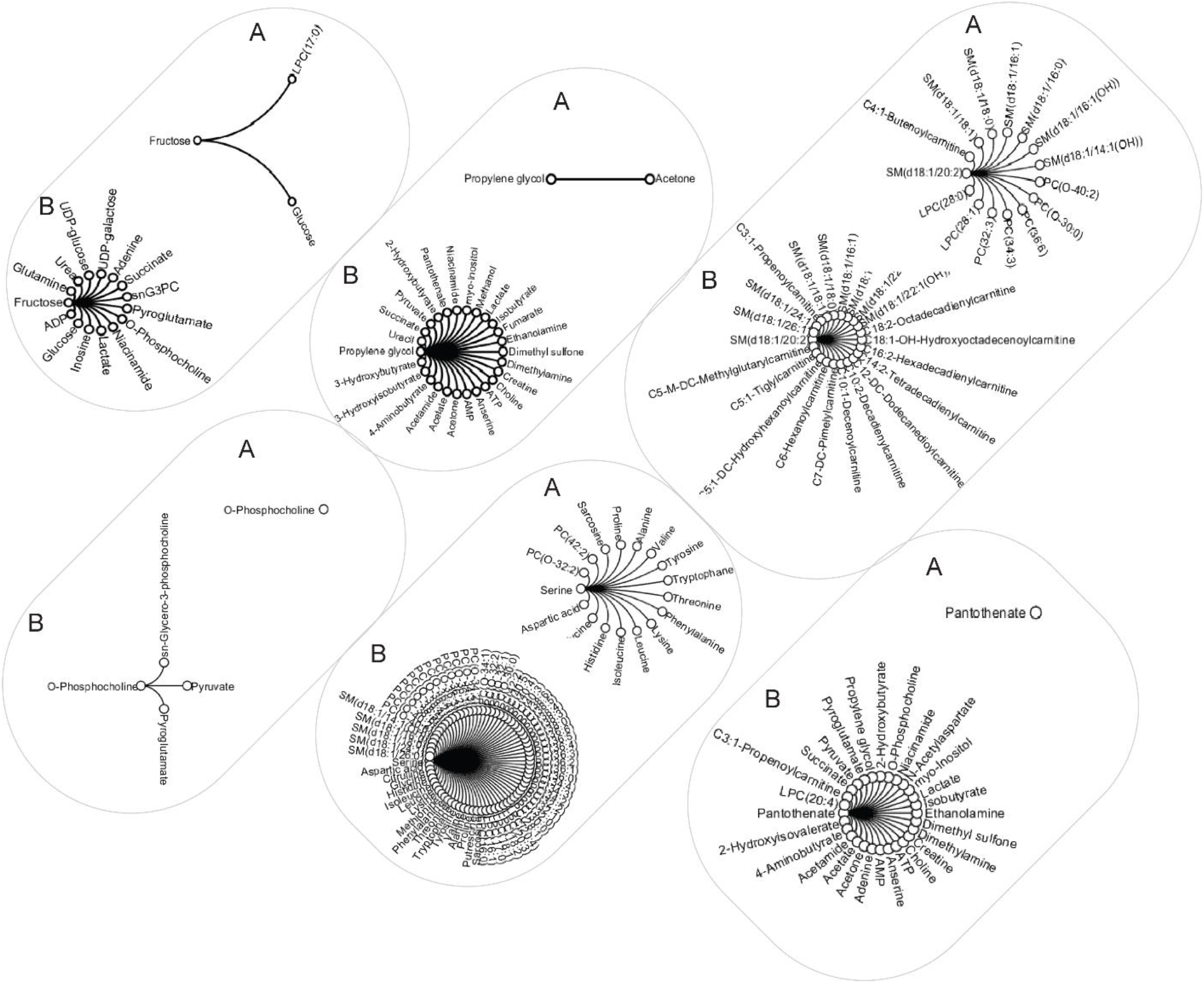
Closest correlation network partners determined using distance correlation for metabolites showing largest overall change in correlation networks. Shown are edges with distance correlation over 0.6 and p-value <0.05 for A. control and B. DLB groups for Fructose, Propylene glycol, SM(d18:1/20:2), O-phosphocholine, serine and Panthothenate. All shown correlations are positive according to Pearson sign derivation.

## 4 Discussion

Network analysis is used to visualize and interpret data and in this work we are showing novel way to use distance correlation network analysis to determine features with significant activity changes. In order to obtain information about both linear and non-linear correlations possibly on data with not strictly normal distribution and importantly provide measure that correlation is only zero when variables are fully independent (Edelmann, et al., 2021) we are introducing application of distance correlation as a method of choice for correlation analysis in metabolomics and lipidomics. Additionally, by combining distance correlation value with the sign of Pearson correlation we introduce a way to determine signed distance correlation for metabolomics data. Application of these approaches is presented on the metabolomics and lipidomics profile of DLB patient brain samples with age and sex matched controls leading to novel information about major metabolic changes in this disease.

Objective diagnosis as well as disease-modifying therapies are lacking for DLB. This progressive and terminal disease is currently only diagnosed based on its clinical presentation, as there are no known biomarkers for early, sensitive and specific detection. Earlier diagnosis and proper stratification would allow for the development and utilization of specific and directed therapies based on an identified and mechanistically-understood metabolic dysregulation. Small-molecule metabolites and lipids are at the intersection of a cell or tissue’s genetic background and environmental history. In particular, the level of a specific metabolite or lipid within a cell or tissue is a consequence of its physiological, developmental, and pathological state (Fiehn, 2002). These levels can be used as a reflection of specific phenotypes (Shamim, et al., 2018). Due to major inter-dependences within the metabolome, only looking at the concentration of each metabolite may be short-sighted. Analysis of the metabolic network differences between patient groups is needed in order to determine functional changes between metabolites and across metabolic pathways and networks. Feature selection through statistical or machine learning approaches focuses on the determination of features that are most relevant for sample classification without considering functional changes that are leading to the observed concentration differences. The development of interaction networks is a major step in understanding the role of metabolites in the pathophysiology of any given disease and the relevance of metabolic markers for diagnosis as well as therapy. Data-driven metabolic networks can be obtained through correlation or classification methods. Small sample size, incomplete knowledge of all the steps in a metabolic pathway, as well as sparse coverage of metabolites present a unique challenge for network derivation. Here, we present metabolic networks for metabolites and lipids in the brain of DLB patients and matching controls as determined using distance correlation analysis. Our analysis revealed major changes in the metabolism of fructose, *O*-phosphocholine, propylene-glycol, SM(d18:1/20:2) and pantothenate (Figure 5). Additionally, number of pairwise correlation changes are observed with for example major changes in serine and proline correlation with members of the phosphcholine family (Figure 4). This is further compounded by the concentration changes for *O*-phosphocholine, *sn*-glycero-3-phosphocholine, putrescine, PC(O-34:0) and hydroxyvalerylcarnitine (C5-OH (C3-DC-M)) and major overall changes in network topology for a number of metabolic families in DLB.

Notably, the correlation network for fructose had significant correlations only with glucose and LPC(17:0) in the control group, but presents many significant edges in the DLB group. The role of fructose and its changing metabolism has been previously hypothesized for different types of dementias and is considered a risk factor in this disease family (Johnson, et al., 2020; Xu, et al., 2020). The link between either endogenous or dietary fructose and the induction of the purine degradation pathway was suggested in Alzheimer’s disease (AD) (Johnson, et al., 2022) and we report its strong correlation in DLB using our distance correlation analysis (Figure 7). The purine degradation pathway is induced by fructose through its rapid depletion of cellular ATP levels and activation of AMP deaminase (AMPD), eventually leading to the production of uric acid. Changes in the level of uric acid have been previously linked to the risk of dementia, with low levels associated with Parkinson’s disease (Ellmore, et al., 2020; Johnson, et al., 2022) and high levels of serum uric acid linked to vascular or mixed dementia and to a lesser extent AD (Latourte, et al., 2018).

Distance correlation analysis showed major differences in correlation partners for fructose in patients with DLB compared to healthy controls, including significant correlations with several metabolites that are part of purine metabolism including ADP, adenine, inosine and urea. Significant changes, especially in certain acylcarnitine and LPC levels, have been previously associated with a high fructose diet (Garcia-Esparcia, et al., 2017). A number of metabolites in the fructose network are also part of lactose synthesis and galactose metabolism. These include glucose, UDP glucose, UDP galactose, ADP and fructose. Fructose production from glucose through the polyol pathway has been previously observed in different tissues including brain (Andres-Hernando, et al., 2019). The production of fructose through this pathway stimulates triglyceride and uric acid accumulation and is hypothesized to be a relevant factor in the development of metabolic syndrome. Up-regulation of aldose reductase, a rate-limiting enzyme in this pathway, has been shown in aging and is indicated as a response to a number of known dementia risk factors (Johnson, et al., 2020). It is important to point out that the concentration of fructose does not show a significant difference between control and DLB cohorts, suggesting a significant change in the fructose metabolic production and utilization rather than accumulation or depletion of this metabolite in these patients.

In both control and DLB cohorts, propylene glycol has a strong correlation with acetone which suggests a link via propanoate metabolism in agreement with previous work linking this pathway to aging and AD (Killingsworth, et al., 2020). Propylene glycol is metabolized to lactate, acetate, and pyruvate and significant correlations with these metabolites exist in the DLB group. In fact, propylene glycol as well as several of its correlation partners, i.e., succinate, acetate, lactate, acetone, pyruvate, 2hydroxybutyrate are involved in propanoate metabolism (based on KEGG map00640 pathway list). The propylene glycol and propanoate metabolism are further linked through the β-Alanine metabolic pathway, which includes anserine (part of β-Alanine metabolism) and pantothenate, both of which correlated strongly with propylene-glycol in DLB. Several metabolites from the propanoate pathway have previously been reported to be significantly different in the saliva of dementia patients (Figueira, et al., 2016) and alterations to this pathway have been observed in AD (Kong, et al., 2014). It is important to point out that propylene-glycol is also used as a solvent for several intravenously administered drugs including lorazepam and diazepam, both of which are used to treat symptoms associated with DLB. The correlation with therapeutic intervention needs further examination, particularly as changes in concentration of pantothenate (vitamin B5), anserine, and dimethyl sulfone could also occur as a result of treatment with either as drugs or supplements (Ding, et al., 2018). For example, anserine is a supplement used to improve symptoms of dementia; dimethyl sulfone is an anti-pain, inflammatory and osteoarthritis drug (Drug Bank ID: DB14090 (Wishart, et al., 2018) and pantothenate is a recommended supplement for dementia patients.

Pantothenate shows one of most significant network changes in the DLB group compared to the control group, with strong correlations to propylene glycol, anserine and dimethyl sulfone. As mentioned previously. pantothenate plays a role in β-alanine metabolism and CoA biosynthesis. In the control group, pantothenate has no significant partners at correlation levels over 0.6 and p-values below 0.01. However, in the DLB group, pantothenate gains several strong correlations, including 4-aminobutanoate, a by-product of one of the steps of β-alanine synthesis, as well as anserine, suggesting its role in metabolism changes in DLB patients.

There are several strengths of the present study. These include the standardized brain collections, the use of age- and sex-matched controls of this unique cohort, a combined quantitative metabolomic approach delivering the most comprehensive metabolite coverage of the DLB brain metabolome, and implementation of novel, analytical approaches within the realm of DLB metabolomics. The small sample size of this unique cohort is a major limitation; however, one should be cognizant of the problems in obtaining such a specialized and well-characterized post-mortem brain sample set. Through the examination of well-characterized samples, even at this sample size, we can develop models of high diagnostic accuracy. It would be worthwhile to expand the analysis carried out here to other brain regions to assess the wider metabolic disturbances in the DLB brain. For any future diagnostic developments, brain tissue is far from an ideal matrix for discovering dementia biomarkers, and it will be necessary to validate our findings in more accessible, non-invasive biological matrices such as blood serum/plasma, as evidenced by Varma et al. for AD (Varma, et al., 2018). Another major limitation which may have provided some additional, useful insight to our exploratory study is the lack of a detailed medical report with important information regarding medications and supplements.

In this study, we introduce a novel approach of signed distance correlation analysis, as well as novel methods for determination of major changes in the correlation network for the study of DLB. The application of these new analytical approaches on such a unique metabolomics data set highlights fructose, propylene-glycol and pantothenate, as well as their associated metabolic pathways as key factors linked with DLB pathology. Novel methods presented in this work are made available for future use for other applications in metabolomics and lipidomics. Our findings have the potential to provide new insight into the pathophysiology of DLB, inspiring development of novel therapeutics and diagnostic methods capable of accurately discriminating DLB from control brain with a high degree of accuracy. Understanding of the major changes in brain metabolism/biochemistry are crucial for future development of treatments.

## Data Availability

All data produced are available online at

https://doi.org/10.4224/40002692

## Data Availability

All data is available at: https://doi.org/10.4224/40002692

## Institutional Review Board Statement

This study was approved by the Beaumont Health System’s Human Investigation Committee (HIC No.: 2018-387).

## Informed Consent Statement

Not applicable.

## Author Contributions

Conceptualization, M.C.C., S.F.G., S.A.L.B.; Methodology, M.C.C., A.Y., S.A., S.A.L.B. and S.F.G.; Formal Analysis, M.C.C.; Experimental Analysis: A.Y.; S.A.; S.V.; S.F.G.; Clinical Samples: B.M.; P.P.; P.G.K.; M.E.M.; B.D.G.; Software development, D.S., A.S. and M.C.C.; Supervision, S.F.G., M.C.C., S.A.L.B.; Writing—Original Draft Preparation, M.C.C; Writing—Review and Editing, M.C.C., S.F.G. I.A., T.N-T.; B.D.G.; A.Y.; All authors have read and agreed to the published version of the manuscript.

## Acknowledgements

We thank the Brains for Dementia Research (BDR) initiative, a brain bank network funded by Alzheimer’s Brain Bank UK (ABBUK), a charity cofunded by Alzheimer’s Research UK (ARUK), and the Alzheimer’s Society, for graciously providing the tissue samples for this study. This work was partly funded by the generous contribution made by the John and Marilyn Bishop Charitable Foundation and the Fred A. & Barbara M. Erb Foundation. Most importantly, we would like to thank all patients and volunteers for their invaluable and selfless contribution to this research.

---

**Supplementary Figure 1**. Comparison of Pearson correlation network between CTRL and DLB using Fisher z-transformation analysis of individual correlation differences. (A) Percentage of correlation values that have statistically significant difference between Pearson and Distance correlations (p<0.05) for control and DLB groups separately. (B) Comparison between Pearson and Distance correlation values for several examples of metabolites with highest percentage of different values. Indicated are metabolites with highest difference between Pearson and Distance correlations in this examples.

